# REECAP: Contrastive learning of retinal aging reveals genetic loci linking morphology to eye disease

**DOI:** 10.1101/2025.11.19.25340555

**Authors:** Liubov Shilova, Daniel Sens, Ayshan Aliyeva, Shubham Chaudhary, Qiaohan Xu, Emmanuelle Salin, Johannes Schiefelbein, Ben Asani, Oana Veronica Amarie, Elida Schneltzer, Ayellet V. Segrè, Julia A. Schnabel, Na Cai, Bjoern M. Eskofier, Francesco Paolo Casale

**Affiliations:** Institute of AI for Health, Helmholtz Zentrum München – German Research Center for Environmental Health, Neuherberg, Germany; Helmholtz Pioneer Campus, Helmholtz Zentrum München – German Research Center for Environmental Health, Neuherberg, Germany; Friedrich-Alexander-Universität Erlangen-Nürnberg, Erlangen, Germany; School of Computation, Information and Technology, Technical University of Munich, Garching, Germany; Department of Ophthalmology, University Hospital, LMU Munich, Munich, Germany; Institute of Experimental Genetics, Helmholtz Zentrum München, Munich, Germany; Department of Ophthalmology, Massachusetts Eye and Ear, Harvard Medical School, Boston, MA, USA; Broad Institute of Harvard and MIT, Cambridge, MA, USA; Institute of Machine Learning in Biomedical Imaging, Helmholtz Zentrum München – German Research Center for Environmental Health, Neuherberg, Germany; School of Biomedical Engineering and Imaging Sciences, King’s College London, London, UK; Department of Biosystems Science and Engineering, ETH Zürich, Basel, Switzerland; Institute of AI in Medicine, LMU Klinikum, Munich, Germany

## Abstract

Deep learning foundation models excel at disease prediction from medical images, yet their potential to bridge tissue morphology with the genetic architecture of disease remains underexplored. Here, we present REECAP (Representation learning for Eye Embedding Contrastive Age Phenotypes), a framework that fine-tunes the RETFound retinal foundation model using a contrastive objective guided by chronological age. Applied to 87,478 fundus images from 52,742 UK Biobank participants, REECAP aligns image representations along the aging axis, yielding multivariate ageing phenotypes for genome-wide association studies (GWAS). GWAS of REECAP embeddings identifies 178 loci, including 27 that colocalize with risk loci of age-related eye diseases, 14 of which remained undetected by conventional disease-label GWAS. By enabling conditional image synthesis, REECAP further links genetic variation to interpretable anatomical changes. Benchmarking against alternative embedding models, we show that REECAP enhances both locus discovery and disease relevance of genetic associations, suggesting that aging-informed tissue embeddings represent a powerful intermediate phenotype to discover and interpret disease loci.

## Introduction

Genome-wide association studies (GWAS) have mapped thousands of disease-associated loci, yet translating these signals into tissue-specific biological mechanisms remains a central challenge in human genetics.

One strategy to bridge this gap is to incorporate imaging-derived phenotypes as intermediate traits, which can increase both the statistical power and interpretability of genetic analyses^1–5^. Recent advances in deep learning–based phenotyping have extended this approach^6–13^, yet most efforts remain task-specific and have not been systematically evaluated for their impact on genetic discovery.

Foundation models trained via self-supervised learning on large image datasets and fine-tuned for downstream tasks provide a powerful basis for studying tissue architecture and its links to disease, and are increasingly used for clinical prediction^14–20^. For example, in ophthalmology, the RETFound model achieves state-of-the-art performance in disease classification^20,21^. However, the application of foundation models to genetic discovery remains unexplored.

Here, we introduce REECAP (Representation learning for Eye Embedding Contrastive Age Phenotypes), a framework that fine-tunes the RETFound encoder using a contrastive objective guided by chronological age to capture representations enriched for age-related disease biology. Applied to fundus photographs from the UK Biobank (UKB), REECAP improves genetic discovery and interpretability compared to expert-defined and deep learning–based phenotypes, with enhanced colocalization to age-related eye diseases and links to retinal morphology.

## Results

### Contrastive fine-tuning of foundation models for tissue aging

To enhance the value of image-derived representations for disease genetics, we developed REECAP, a framework that fine-tunes vision foundation models via contrastive learning guided by chronological age, aligning the latent space along a continuous trajectory of retinal aging (**Fig. 1a**). Starting from RETFound^20,21^, we applied REECAP to 87,478 high-quality fundus images from 52,742 UKB participants and used the resulting embeddings as multivariate phenotypes in a GWAS (**Supplementary Fig. 1**).

**Figure 1.**
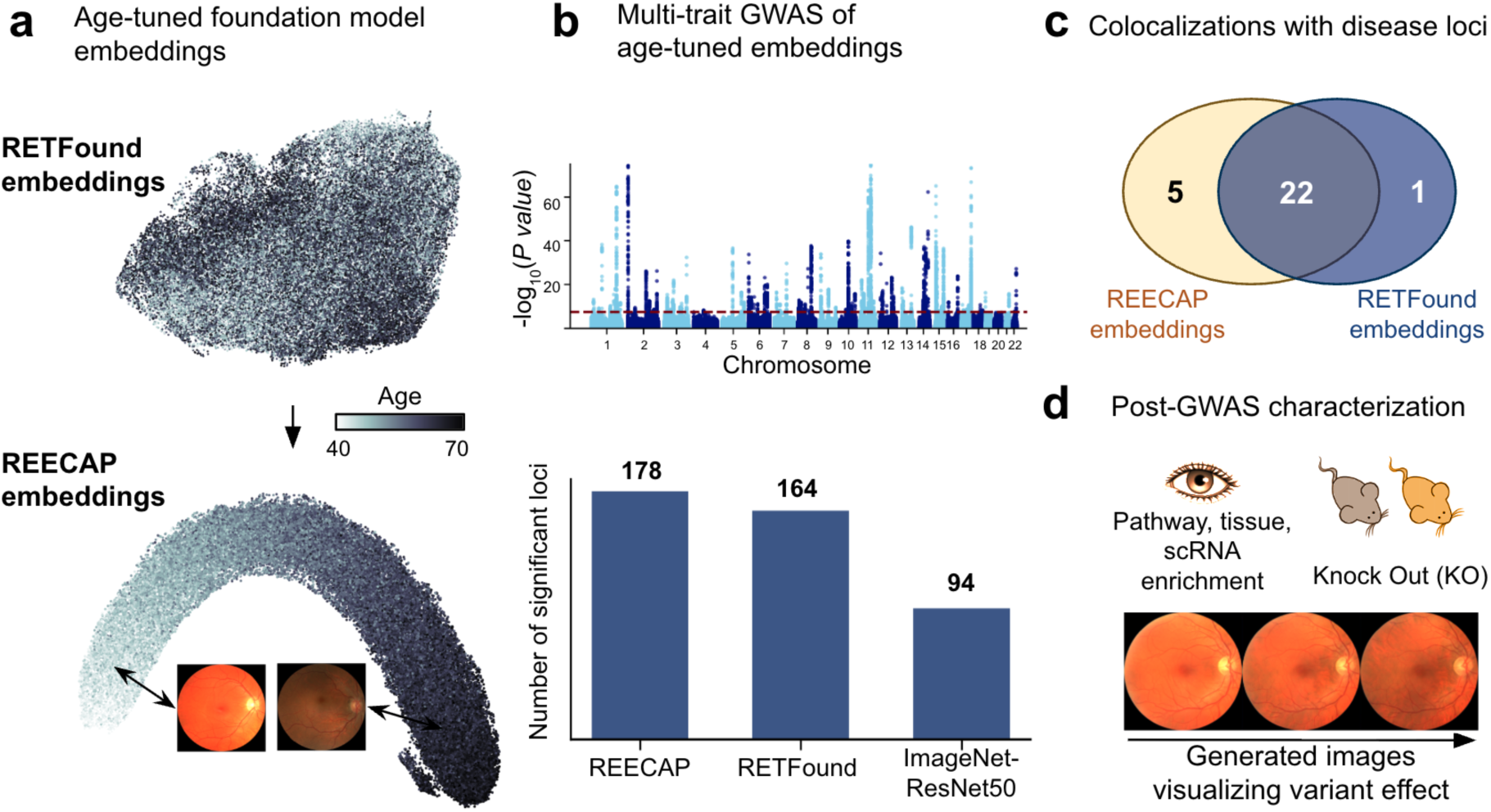
Overview of the REECAP framework. (**a**) UMAP projection of fundus embeddings from RETFound (top) and REECAP (bottom). Contrastive fine-tuning on chronological age aligns the latent space along a continuous retinal aging axis, illustrated by example images from opposite extremes. (**b**) Multi-trait GWAS of REECAP embeddings identifies more genome-wide significant loci than baseline models. Top: Manhattan plot of REECAP associations. Bottom: number of loci detected by REECAP, RETFound, and ResNet50 (ImageNet-pretrained). (**c**) Disease relevance of REECAP loci. Venn diagram of colocalizations with ocular disease loci from FinnGen using REECAP versus RETFound embeddings. (**d**) Downstream characterization of REECAP loci, including pathway, tissue, and cell-type enrichment; conditional image synthesis of retinal morphology; and overlap with in vivo mouse knockout data.

To determine whether REECAP embeddings provide more informative signals for disease genetics than other image-derived phenotypes, we benchmarked them against alternative multivariate and univariate approaches (**Fig. 1b**). Beyond discovery, we assessed disease relevance through colocalization with major ocular disease GWAS from FinnGen (**Fig. 1c**), as well as polygenic risk prediction and pathway enrichment.

We next examined the biological processes and loci captured by REECAP (**Fig. 1d**). Specifically, we linked both known and novel disease loci discovered through REECAP to morphological changes in the eye using conditional image generation^12^, producing synthetic fundus images conditional on REECAP embeddings, and to functional evidence from mouse knockout databases.

### Age-tuned embeddings capture biologically relevant retinal features

To capture eye features aligned with aging, we fine-tuned the RETFound encoder using a contrastive objective based on chronological age^22^, which encourages representations of patients with similar ages to be closer than those of patients with larger age differences, effectively organizing the latent space along a continuous age trajectory. We applied this training to 87,478 high-quality fundus images from 52,742 UKB participants (**Methods**). The resulting REECAP representations substantially improved age prediction by linear probing (mean absolute error [MAE] = 2.86 ± 0.007 years), compared with the original RETFound representations (MAE = 3.40 ± 0.01 years; P < 3×10^-7^; **Fig. 2a**, **Methods**). REECAP also outperformed conventional supervised regression-based fine-tuning (MAE = 2.86 ± 0.007 versus 3.03 ± 0.003 years; P < 5×10^-9^; **Methods**); **Supplementary Fig. 2**), consistent with prior work showing superior performance of contrastive objectives over standard regression losses^22^.

**Figure 2.**
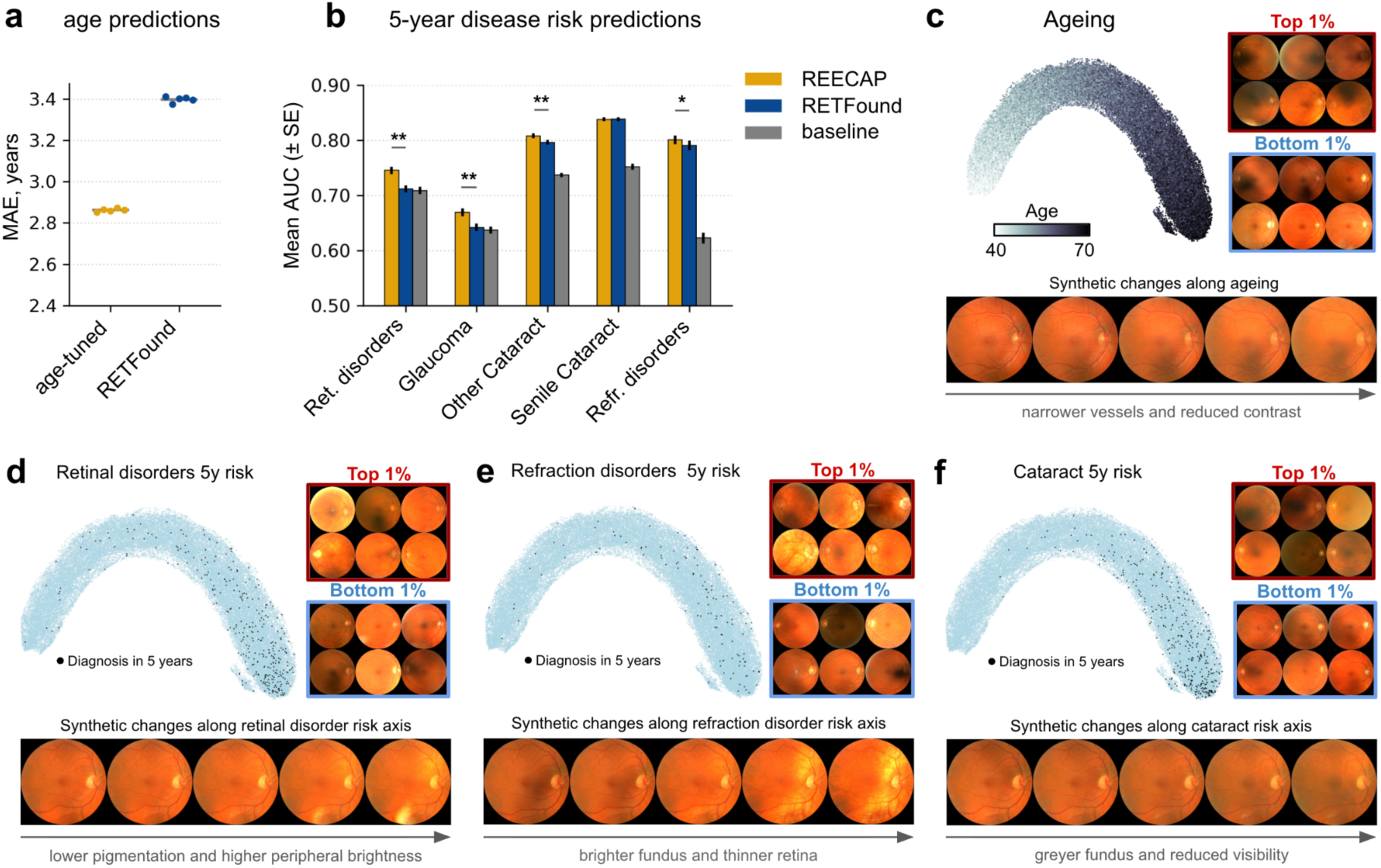
REECAP embeddings improve age prediction and reveal disease-relevant retinal features. (**a**) Mean absolute error (MAE) for chronological age prediction using contrastively fine-tuned (REECAP) versus untuned RETFound embeddings. (**b**) Five-year disease risk prediction using ridge logistic regression trained on REECAP (gold), RETFound (blue), or scalar baseline features (gray). Bars show mean area under the ROC curve (AUC) across 20 random splits; *P < 0.05, **P < 0.0001 (paired t-test with Bonferroni correction). (**c**–**f**) Morphological variation along embedding-derived axes for (**c**) age, (**d**) retinal disorders (ICD10 H35), (**e**) refraction disorders (H52), and (**f**) cataracts (H25). Each panel shows a UMAP of REECAP embeddings colored by trait value, real fundus images from opposite ends of the axis, and synthetic images generated by a conditional generative model (**Methods**).

We next assessed the clinical relevance of REECAP by predicting five-year risk for major ocular diseases. REECAP embeddings improved risk prediction for glaucoma, retinal disorders (including age-related macular degeneration, AMD), and refraction disorders compared to untuned embeddings (Bonferroni-adjusted P < 0.05; **Fig. 2b–d**, **Methods**). Across all five conditions, REECAP performed as well as or better than both untuned RETFound and baseline models based on demographic variables (**Methods**).

To evaluate whether REECAP embeddings capture specific morphological features linked to aging and pathology, we leveraged a conditional generative framework to visualize patterns associated with age and future diagnosis. Specifically, we trained generative adversarial networks to generate synthetic images conditional on REECAP embeddings, using interpolation to visualize fundus changes across gradients of age and five-year disease risk (**Methods**, **Supplementary Fig. 3-7**)^12^. The resulting synthetic images were reviewed by two ophthalmologists, who confirmed distinct, clinically relevant patterns along each phenotypic axis. For aging, reconstructions showed progressive vessel narrowing and reduced image contrast, consistent with healthy fundus aging^23–25^ (**Fig. 2c**, **Supplementary Fig. 3**). For retinal disorders, higher risk was associated with a slightly less pigmented fundus (**Fig. 2d**), consistent with early hypopigmentation of the retinal pigment epithelium—a known prognostic marker for progression to late AMD^26^—with individual samples additionally showing peripheral atrophy, pale optic discs, and altered foveal reflexes (**Supplementary Fig. 4**). Refraction disorder risk was linked to myopic features, including a significantly brighter fundus and an atrophic-appearing, thinner retina (**Fig. 2e, Supplementary Fig. 5**). Along the cataract risk axis, images appeared greyer with reduced fundus visibility and subtle vessel thinning, consistent with cataract-related media opacity^27^ (**Fig. 2f**, **Supplementary Fig. 6**).

Together, these analyses show that REECAP embeddings improve disease risk prediction and enable biological interpretation through generative modeling, capturing morphological features aligned with aging and ocular pathology.

### GWAS of REECAP embeddings identifies loci and pathways linked to eye aging and disease

To dissect the genetic architecture of eye aging captured by REECAP, we performed a multi-trait genome-wide association study (GWAS) using all embedding dimensions as a joint phenotype. The analysis included 36,349 unrelated European UKB participants with genotype and high-quality imaging data. We adjusted for age, sex, and the top 20 genetic principal components to account for population structure (**Methods**). Across 9,447,684 analyzed common variants (minor allele frequency ≥1%), we identified 178 independent genome-wide significant loci (P < 5×10⁻⁸; **Fig. 3a**, **Methods**), with well-calibrated test statistics under permutation-based null models (**Supplementary Fig. 8**, **Methods**). This multivariate approach nearly doubled the number of discovered loci compared to a univariate GWAS on each embedding dimension with Cauchy-based P value aggregation^28^ (178 vs. 90 loci; **Supplementary Fig. 9**, **Methods**).

**Figure 3.**
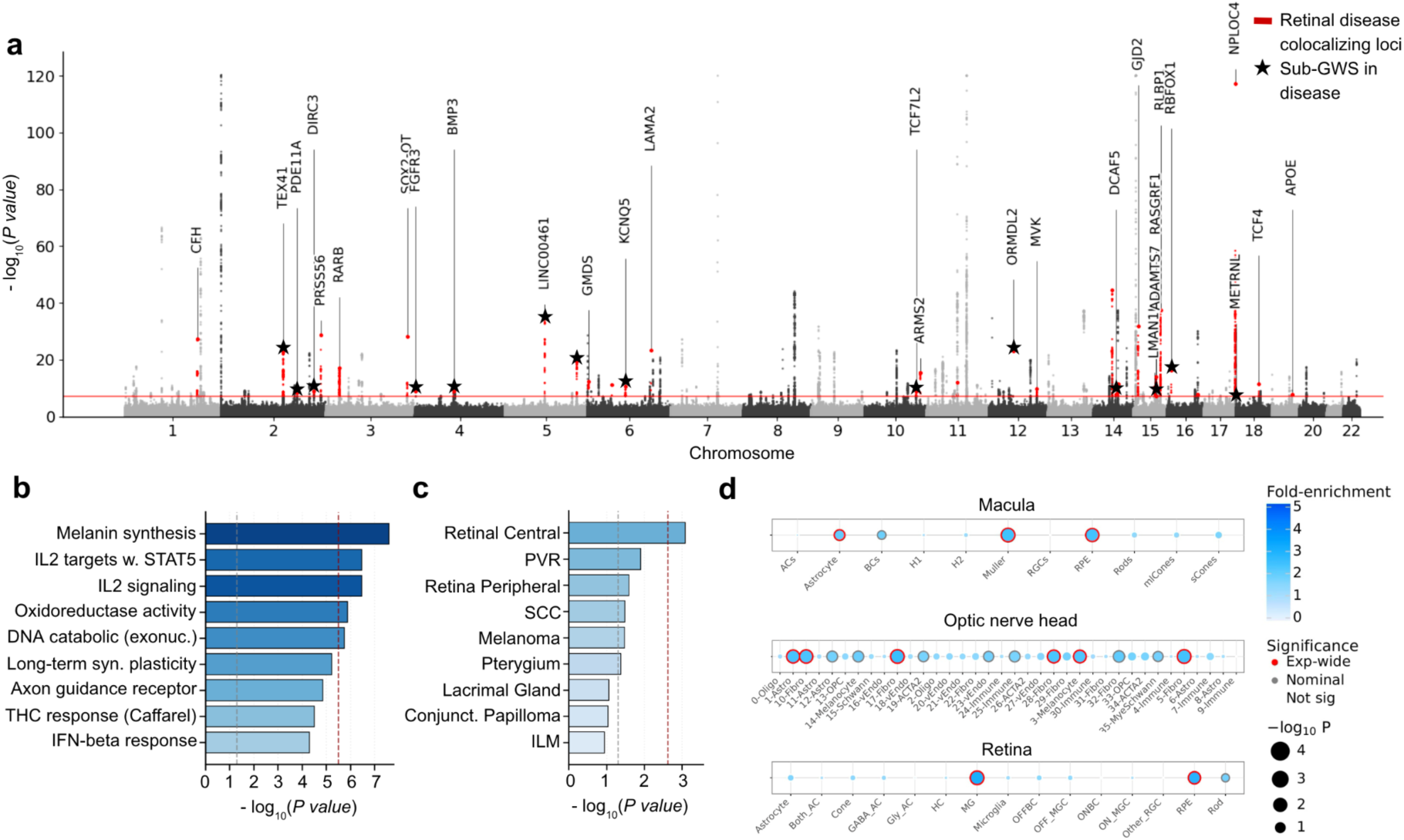
Multi-trait GWAS of REECAP embeddings identifies loci, pathways, and cell types linked to retinal aging. (**a**) Manhattan plot of multi-trait GWAS of REECAP embeddings. Red points indicate loci colocalizing with ocular disease GWAS signals in FinnGen (PP-H4 > 0.8); black stars denote loci not genome-wide significant in FinnGen alone. Annotated genes mark closest genes to lead variants. (**b**) MAGMA gene-set enrichment highlights pigmentation, immune signaling, oxidative stress, and DNA damage response pathways. Bars show –log₁₀(P); red dashed line indicates Bonferroni threshold, black dashed line indicates nominal P = 0.05. Colors represent biological categories. (**c**) MAGMA tissue enrichment identifies the central retina as the top enriched tissue. Bars represent –log₁₀(P); significance thresholds as in **b**. (**d**) ECLIPSER analysis of cell-type enrichment in single-nucleus RNA-seq from three ocular tissues. Bubble size reflects –log₁₀(P), color denotes fold enrichment, and border shading indicates significant enrichments (5% FDR).

Colocalization with intermediate traits revealed that 41 of 178 loci (24%) overlap with retinal vessel density^29^ and 37 (21%) with hair color^30^ (PP-H4 > 0.8; **Methods**, **Supplementary Dataset 1**), highlighting vascular and pigmentation pathways as contributors to retinal aging.

Functional interpretation of common variant signals highlighted involved biological pathways. MAGMA enrichment across 17,023 MSigDB gene sets revealed 5 significantly enriched pathways (Bonferroni-adjusted P < 2.94×10^-6^, **Fig. 3b**, **Methods**), implicating pigmentation, immune regulation, oxidative stress, and DNA damage responses as central processes in eye aging. Additional non-significant enrichments pointed to synaptic plasticity and axon guidance, consistent with neurodegenerative mechanisms in eye disease^31–33^.

Tissue- and cell-type enrichment analyses based on common variant signals supported a retinal origin for these signals, with the central retina emerging as the top-enriched tissue (**Fig. 3c**, **Methods**). Within the retina and macula, enrichment of genes mapped to the GWAS loci was strongest in astrocytes, Müller cells, and retina pigment epithelium (RPE) (FDR < 5%, **Fig. 3d**, **Methods**, **Supplementary Fig. 10**), while in the optic nerve head, astrocytes and fibroblasts were also implicated (FDR < 5%), consistent with recent reports^34,35^.

Integration with ocular disease GWAS in FinnGen revealed convergence with ocular disease. Of the 178 loci, 27 (15.2%) colocalized with at least one eye disease (PP-H4 > 0.8, **Methods**), including 14 loci not reaching genome-wide significance in FinnGen alone (**Fig. 3a**, black stars; **Supplementary Dataset 1**). Colocalizations were most frequent with cataract, followed by myopia, AMD, and glaucoma, with 9 loci colocalizing with more than one disease. In comparison, the univariate GWAS yielded only 12 disease-colocalizing loci (**Methods**, **Supplementary Fig. 11**).

Finally, applying the same multivariate framework to gene-level burden scores from rare coding variants in 17,984 genes within UKB revealed significant associations in melanin biosynthesis enzymes (*TYR*, *OCA2*, *SLC45A2*, *TYRP1*), visual cycle components (*RLBP1*, *RAB32*), and the matricellular protein *THBS4*, a regulator of vascular remodeling and age-related expression (**Supplementary Fig. 12**, **Methods**).

Collectively, these results position REECAP as a framework for linking image-derived phenotypes to genetic variation, revealing pathways and cell types relevant to eye aging and disease.

### REECAP enhances genetic discovery and disease relevance over baseline models

To assess whether REECAP embeddings provided more informative genetic signals than other image-derived phenotypes, we benchmarked them against alternative representations: embeddings from the untuned RETFound encoder, embeddings from a domain-agnostic ResNet50 model (as in transferGWAS^36^), a scalar retinal age predictor, and the canonical imaging trait cup-to-disc ratio. We compared each method on the number of loci discovered, colocalization with disease GWAS, polygenic risk prediction, and pathway enrichment.

REECAP embeddings identified the largest number of independent genome-wide significant loci (178) and disease colocalizations (27), outperforming untuned RETFound (164 loci, 23 colocalizing), ImageNet-ResNet50 (94, 11), cup-to-disc ratio (26, 5), and scalar retinal age (6, 1) (**Fig. 4a–b, Methods**, **Supplementary Fig. 9, 11, 13**). Genetic risk scores derived from genome-wide significant loci were trained and validated on multiple UK Biobank train/test splits, with REECAP-based scores consistently outperforming baseline models in disease prediction accuracy (**Fig. 4c**; **Methods**).

**Figure 4.**
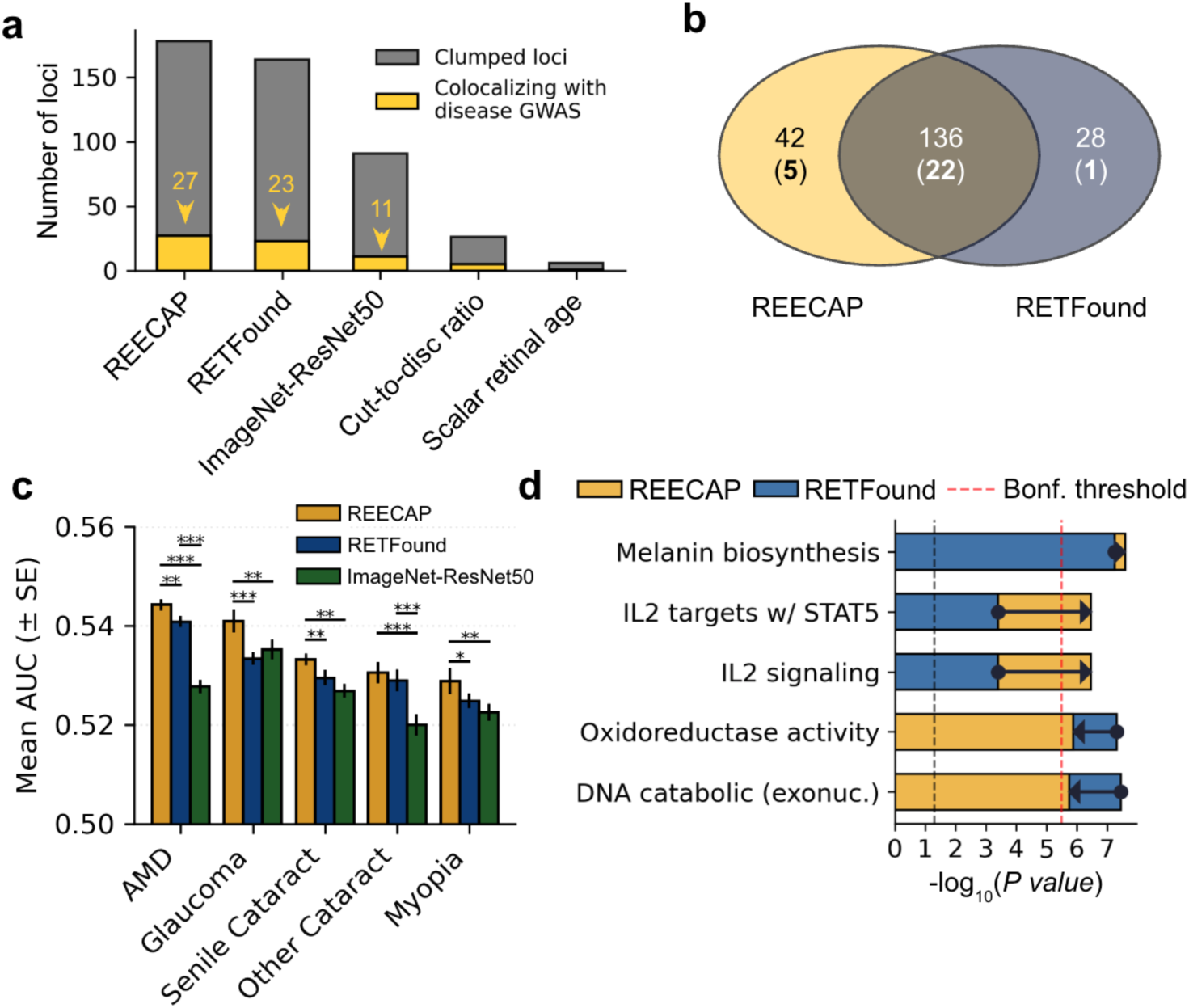
REECAP embeddings outperform baseline image-derived phenotypes across genetic and disease-relevance benchmarks. (**a**) Number of independent genome-wide significant loci (gray) and disease GWAS colocalizations (PP-H4 > 0.80; yellow) identified using five imaging-derived traits: REECAP, RETFound, ImageNet-ResNet50, cup-to-disc ratio, and scalar retinal age. (**b**) Overlap between loci detected using REECAP versus RETFound embeddings; numbers in parentheses indicate loci colocalizing with disease GWAS. (**c**) Mean area under the ROC curve (AUC ± s.e.m.) for polygenic risk score (PRS) models of five ocular diseases, constructed using independent variants from each embedding type. *P < 0.05, **P < 0.01, ***P < 0.001 (paired t-test, N = 10). (**d**) MAGMA pathway enrichment (–log₁₀P) for REECAP (yellow) versus RETFound (blue) across selected MSigDB gene sets. Arrows indicate differences between models. Red and black dashed lines denote Bonferroni and nominal significance thresholds, respectively.

Pathway analyses confirmed the disease relevance of REECAP signals. Although REECAP and RETFound embeddings exhibited similar heritability (**Methods**, **Supplementary Fig. 14**), REECAP prioritized pathways central to retinal biology, including melanin biosynthesis, *IL-2/STAT5* signaling, and oxidoreductase activity, while deprioritizing oxidoreductase and DNA catabolic programs strongly enriched in RETFound embeddings (**Fig. 4d**). In contrast, ImageNet-ResNet50 signals were enriched for more generic transcriptional pathways with limited connection to ocular aging (**Supplementary Dataset 2**).

Together, these findings show that REECAP embeddings yield genetic signals with stronger biological and disease relevance than either generic, unsupervised, or expert-defined image features, establishing fine-tuned representation learning as a powerful strategy for image–genomic phenotyping.

### Biological interpretation of REECAP loci across morphology and function

Among the 27 loci colocalizing with ocular disease, 13 mapped to well-established risk regions, providing biological validation of the REECAP approach. At *CFH*, the intronic variant *rs10737680* (P-REECAP < 10^-25^) colocalized with four disease outcomes, including AMD and cataract (**Supplementary Dataset 1**), consistent with *CFH* role as a regulator of the complement cascade in retinal inflammation and aging^37–46^. Similarly, *rs3750846* at the *ARMS2–HTRA1* locus (P-REECAP < 10^-15^) colocalized with all analyzed FinnGen eye outcomes (**Fig. 5a**, **Supplementary Dataset 1**) and has been extensively linked to AMD^47–55^. Synthetic image reconstruction revealed progressive fundus hypopigmentation along the risk allele trajectory (**Fig. 5a**), consistent with early retinal pigment epithelium (RPE) thinning and melanin loss.

**Figure 5.**
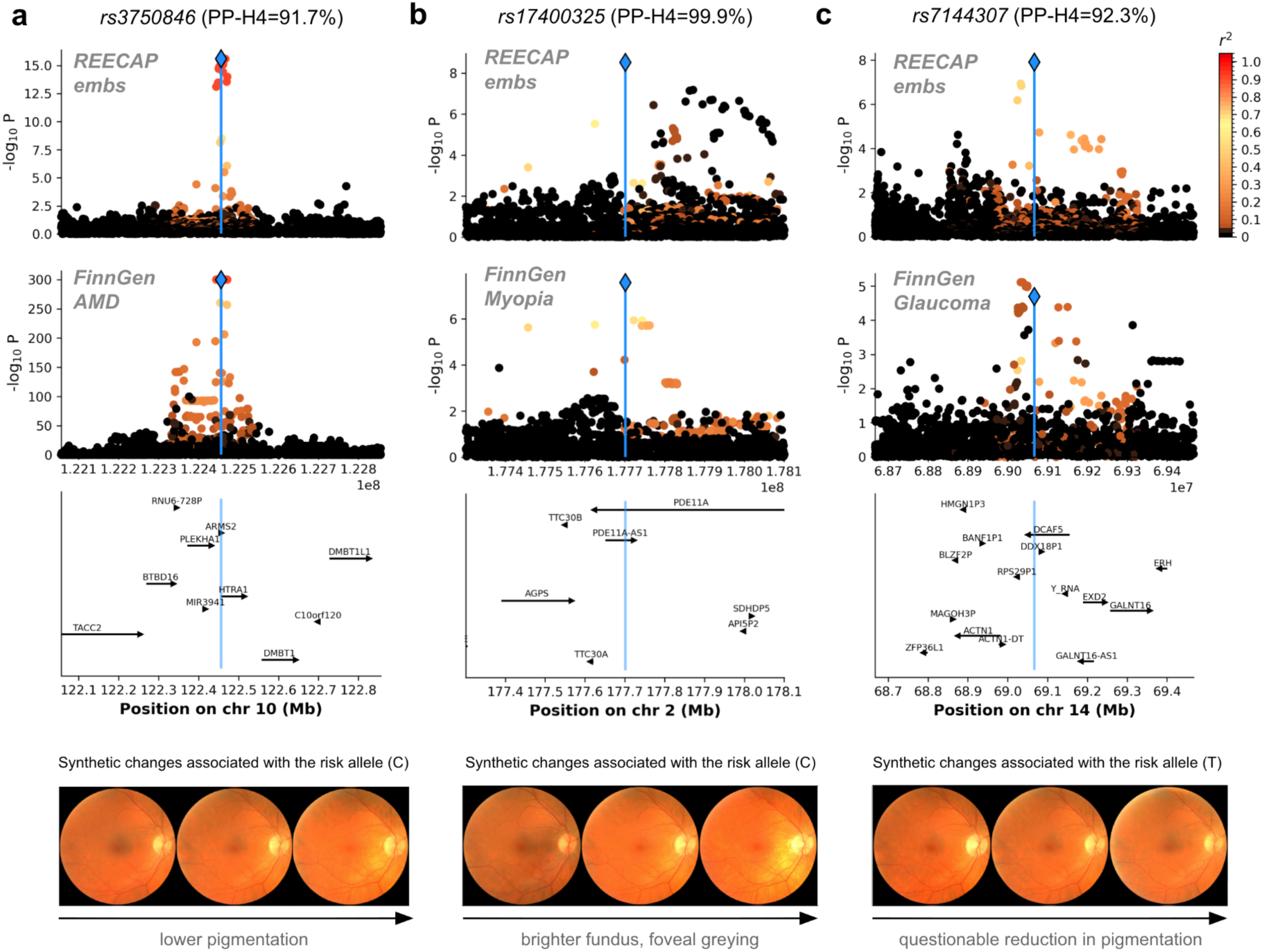
Genetic loci identified by REECAP colocalize with ocular disease and enable morphological interpretation. (**a**–**c**) Representative loci colocalizing with ocular disease in FinnGen: *rs3750846* (*HTRA1–ARMS2*), *rs17400325* (*PDE11A*), and *rs7144307* (*DCAF5*). For each locus, shown are: (i) locus-specific Manhattan plot from the REECAP multi-trait GWAS, (ii) corresponding association signal from the FinnGen disease GWAS, (iii) gene annotations across the region, and (iv) synthetic fundus images generated by a conditional generative network, illustrating predicted morphological changes associated with the risk allele. Risk alleles are defined based on FinnGen disease associations (**Methods**).

Beyond these established loci, REECAP uncovered 14 additional colocalizations not detected at genome-wide significance in disease-specific GWAS. Several signals reinforced prior links to ocular health: association signals at *KCNQ5* colocalized with myopia (P < 10^-10^; PP-H4=0.996; **Supplementary Fig. 15**) and have been implicated in refractive error^56^; *BMP3* signals colocalized with retinal detachment (P-REECAP < 10^-8^; PP-H4=0.906; **Supplementary Fig. 15**), consistent with previous evidence^57^; and a locus in *TCF7L2*, widely associated with diabetes^58–60^, colocalized here with retinal disorders, including diabetic retinopathy (P-REECAP < 10^-8^; PP-H4=0.993; **Supplementary Fig. 15**). A locus near *FGFR3* colocalized with cataract, in line with *FGFR3*’s established role in cataractogenesis^61,62^.

REECAP additionally revealed loci with limited prior evidence linking them to ocular phenotypes. At *PDE11A*, the missense variant *rs17400325* (*Y727C,* P-REECAP < 3×10^-9^) colocalized with myopia in FinnGen (PP-H4 = 0.999; **Fig. 5b**) despite not reaching genome-wide significance there. Rare-variant analyses have previously implicated *PDE11A* in refractive error and myopia^63,64^, and conditional image generation linked the risk allele to a brighter fundus and foveal greying, consistent with choroidal thinning in myopic remodeling^65^. Similarly, variant *rs7144307* in *DCAF5* (P-REECAP < 1.2×10^-8^) strongly colocalized with glaucoma (PP-H4 = 0.923; **Fig. 5c**) despite being subsignificant in the disease-label GWAS. *DCAF5* is expressed in the RPE, microglia, and optic nerve^66^, and while conditional image generation suggested only subtle reduction in retinal pigmentation (**Fig. 5c**), knockout mice (*Dcaf5^em1(IMPC)Ccpcz^* allele) display abnormal retinal morphology, including dysplasia and retinal thinning, supporting a role in retinal homeostasis^67^. Other loci with moderate prior links to eye disease included variants in *RBFOX1, TEX41, LINC00461, ORDML2,* and *METRNL* (**Supplementary Fig. 15**).

Together, these analyses show that REECAP identifies both established and previously overlooked genetic associations and provides biological interpretation by linking risk variants to disease associations and retinal morphology.

## Discussion

We introduce REECAP, an age-guided contrastive learning framework that fine-tunes vision foundation models to generate tissue ageing representations for genetic discovery, extending current univariate approaches to study tissue aging^68–70^. Applied to 87,478 fundus images from 52,742 individuals in the UK Biobank, REECAP identified 178 genome-wide significant loci, 68 of which (38%) colocalized with vessel density and pigmentation traits, supporting vascular and melanocytic changes as hallmarks of retinal aging. Enrichment analyses further implicated oxidative stress, immune regulation, and DNA damage pathways, with retinal glia and pigment epithelium emerging as key cellular contributors.

Beyond locus discovery, REECAP genetic signals act as priors for disease genetics, with 27 of the 178 loci colocalizing with major age-related eye diseases, including 14 missed by conventional disease-label GWAS. To visualize morphological changes associated with risk alleles, we used a conditional generative approach to synthesize fundus images along embedding-derived axes, which we validated on aging and disease-risk trajectories. We applied this framework to risk alleles at both known and novel loci, extending recent generative approaches for modeling genetic and chemical perturbations^12,71,72^.

In a systematic genetics-centered comparison, we show that REECAP outperforms untuned foundation models, domain-agnostic embeddings, and expert-defined features across locus discovery, disease colocalization, and risk prediction. These findings suggest that REECAP can provide a general strategy for tuning vision foundation models for genetic analysis, offering a blueprint for applications to other tissues, modalities, and biological contexts.

REECAP recapitulated key loci involved in retinal aging and disease, including *CFH* and *ARMS2–HTRA1*, supporting its biological validity. It also uncovered novel associations: a missense variant in *PDE11A* (*rs17400325*) colocalized with myopia risk, consistent with rare-variant studies of refractive error^63,64^. Another locus, *DCAF5*, colocalized with glaucoma despite no prior human association; functional support from mouse knockout models showing retinal abnormalities^67^, together with its role in the CUL4–DDB1 ubiquitin ligase complex^73^, implicates *DCAF5* in retinal proteostasis and aging.

Several limitations warrant consideration. The genetic analyses were limited to individuals of European ancestry, and extending REECAP to more diverse populations will be essential for equitable genetic discovery. While REECAP embeddings capture aging-related variation, they are not explicitly tuned to disease-specific axes, which may be addressed in future domain-adaptive extensions. Finally, although generative reconstructions offer biologically plausible interpretations, empirical validation in model systems such as retinal organoids, animal knockouts, or longitudinal imaging will be necessary to confirm anatomical fidelity.

Together, these findings establish REECAP as a generalizable strategy for tuning vision foundation models for genetic analyses. By linking high-dimensional image features to genetic architecture and back to interpretable anatomy, REECAP advances the resolution, interpretability, and translational potential of imaging-genetic studies and offers a blueprint for foundation model alignment in diverse biomedical domains.

## Methods

### Data and Preprocessing

We analyzed genetic data and fundus photographs from the first imaging visit of 52,742 UK Biobank (UKB) participants^74^. High-quality images were identified using the MCF-Net model, retaining 87,478 images with a rejection probability below 80% ^75^. Genetic analyses were conducted in 36,349 unrelated individuals of European ancestry ^76^. Chronological age was obtained from UKB field 21003, and ICD-10 diagnosis codes were derived from hospital records, general practitioner reports, and self-reported data. Eye-related conditions were mapped using UKB category 2407.

### The REECAP framework

REECAP is a representation learning framework that fine-tunes a foundation vision model to capture age-related morphological variation and trains a generative decoder to invert this embedding space, enabling direct visualization of changes along axes defined by traits or genetic variation. This joint architecture supports both statistical association analyses and interpretable image synthesis. In this study, we applied REECAP to retinal aging, initializing the model with the RETFound encoder, a ViT-large transformer pretrained for fundus image reconstruction ^20,77^.

#### Contrastive learning of aging

To align embeddings with continuous biological variation, we fine-tuned the RETFound encoder using Rank-N-Contrast (RnC), a contrastive learning framework optimized for regression tasks^22^. RnC organizes the latent space by enforcing ordinal relationships among samples within a batch: for each image, the model is trained to identify, among all younger samples, the one whose representation is closest to the focal point in latent space (**Supplementary Information**). This optimization encourages local ordering in accordance with chronological age, thereby structuring embeddings along the aging trajectory. Compared to direct regression losses, this approach yields more biologically informative representations and improves downstream age prediction^22^. We fine-tuned the RETFound model using images from both eyes, ensuring that images from the same participant were assigned to the same split. We evaluated performance via five-fold cross-validation, allocating 72% of the data for training, 8% for early stopping and augmentation selection, and 20% for testing (**Supplementary Fig. 16**). Training was performed using the AdamW optimizer^78^ (learning rate = 5 × 10^-6^; weight decay = 1 × 10^-5^) with a batch size of 64.

#### Embedding definition

REECAP embeddings were obtained by averaging the 1,024-dimensional output of the fine-tuned RETFound encoder from left and right eye, followed by principal component analysis for dimensionality reduction prior to GWAS and visualization. For computational tractability, we retained the top 40 components, a number consistent with prior multivariate GWAS applications^8,12^. For visualization, UMAP was applied to a k-nearest neighbor graph (k = 10) using the `scanpy` package^79^.

#### Generative decoder

To visualize the image-level information encoded by REECAP embeddings, we trained a conditional generative adversarial network (GAN) to synthesize fundus images from the 40-dimensional embeddings following the architecture in HistoGWAS^12^. The generator maps a latent variable **z**, sampled from a normal distribution whose mean and variance are functions of the REECAP embeddings **x**, to a 256 × 256 fundus image. The discriminator receives the embedding together with either a real or a synthetic image and is trained to distinguish between using a combination of conditional and unconditional adversarial losses, within a Wasserstein GAN framework with gradient penalty^80,81^. We also employed progressive growing of both generator and discriminator to enhance stability and image quality^82^. Architectural and training hyperparameters were aligned with those used in HistoGWAS^12^ (**Supplementary Fig. 17**).

#### Conditional image generation

To visualize morphological changes associated with age, disease status, and genetic variation, we used the trained generative model to synthesize fundus images across relevant embedding trajectories^12^. For each trait of interest, we fitted a linear mixed model with the trait as the outcome, REECAP embeddings as random effects, and age, sex, and the top 20 genetic principal components as fixed effects. Leave-one-out (LOO) predictions of the random effect were computed for each individual^83^, and mean embeddings were derived for individuals in the bottom and top 0.1% of the predicted distribution, denoted *x_low_* and *x_high_*, respectively. We then defined interpolated embeddings as:

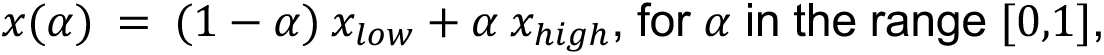

and projected these embeddings back to image space using the GAN generator, enabling visual inspection of morphological changes along the interpolation axis. Multiple synthetic images can be generated for the same interpolated embeddings by sampling distinct latent noise vectors. The full procedure is detailed in **Supplementary Information**.

### Validation of REECAP Embeddings

#### Age prediction performance

We benchmarked the predictive accuracy of REECAP embeddings against both untuned RETFound representations and supervised fine-tuning using a five-fold cross-validation framework. For REECAP and RETFound, ridge regression models with internal cross-validation for hyperparameter tuning were trained to predict chronological age from the corresponding representations. For the supervised baseline, we appended an average pooling layer and a linear output layer to the RETFound encoder and fine-tuned the model end-to-end using mean squared error loss. Training involved 15 warm-up epochs for the linear head (learning rate = 10^-2^), followed by full model optimization at a reduced learning rate (10^-6^). Prediction accuracy was evaluated using mean absolute error (MAE) between predicted and actual age in the held-out folds.

#### Disease risk prediction

We compared the ability of REECAP and untuned RETFound embeddings to predict five-year risk of major ocular diseases. ICD-10 diagnosis codes included other retinal diseases (H35; including age-related macular degeneration), glaucoma (H40), senile cataract (H25), other cataract (H26), and refraction disorders (H52; including myopia). For each method, the top 40 principal components of the representations were used as input features. To benchmark performance, we trained ridge logistic regression models using either REECAP or RETFound embeddings. As an additional baseline, we also evaluated a model using only demographic covariates: age, sex, age^2^, sex × age, and sex × age^2^. Predictive accuracy was measured using area under the receiver operating characteristic curve (AUC), averaged over 20 random 80/20 train–test splits, with five-fold cross-validation in each training set for hyperparameter tuning. Statistical significance of pairwise performance differences was assessed using paired t-tests across the test sets.

#### Visualization of ageing- and disease-associated changes

To assess whether REECAP embeddings capture meaningful morphological variation, we applied the conditional image generation described above to the following traits: age, retinal disorders, including AMD (ICD10 code H35: “other retinal diseases”), cataract (ICD10 code H25), and refraction disorders, including myopia (ICD10 code H52: “disorders of refraction and accommodation“). For each trait, synthetic images were generated by decoding interpolated embeddings along the corresponding direction. Multiple image realizations were produced per interpolation step by sampling different noise vectors. Two ophthalmologists reviewed the generated image sequences along each interpolation and documented characteristic morphological changes.

### Genetic analysis of REECAP embeddings

#### Multi-trait GWAS

To identify genetic associations with REECAP embeddings, we performed a multi-trait genome-wide association study (MT-GWAS) using a re-implementation of the single-variant model described in ^84^. Each embedding vector was modeled as a multivariate phenotype, and genetic effects were tested using a chi-squared statistic with 40 degrees of freedom. Residual trait covariance was estimated once under the null model and only partially optimized under the alternative for faster computation^84–86^. Full model details are provided in the **Supplementary Information**. GWAS was conducted in 36,349 unrelated individuals of European ancestry, testing 9,447,684 variants with minor allele frequency (MAF) ≥ 1%. Covariates included genotyping array, assessment center, age, sex, age^2^, sex × age, sex × age^2^, and the top 20 genetic principal components. Embedding dimensions were rank-normalized to standard Gaussian distributions to mitigate model mismatch due to deviation from Gaussian distribution. As a baseline, we also performed a univariate GWAS by testing each embedding dimension independently and combining P values using the aggregated Cauchy association test^28^.

#### Independent genome-wide loci and colocalization

Independent genome-wide significant loci (P < 5 × 10^-8^) were identified using PLINK’s clumping procedure^87^ with an LD threshold of r^2^ < 0.1 and a 5 Mb window. For each locus, we assessed overlap with disease-associated signals from genome-wide association studies of 10 ocular traits, using summary statistics from FinnGen (**Supplementary Information**). Loci that were genome-wide significant in the REECAP GWAS and associated with at least one ocular trait at a Bonferroni-adjusted P < 0.05 were tested for colocalization using the COLOC method^88^. We additionally evaluated colocalization with GWAS summary statistics for arterial vascular density^29^ and hair color traits^30^ (**Supplementary Information**).

#### Pathway Enrichment Analysis

To interpret biological pathways enriched in the multi-trait GWAS results, we used FUMA with integrated MAGMA^89^ to perform gene-set enrichment analysis under default settings. Variant-level genome-wide association statistics were mapped to genes using MAGMA’s SNP-wise mean model, with a default 0 kb SNP-to-gene window and LD estimated from the European 1000 Genomes Phase 3 reference panel. Enrichment was tested using MAGMA’s competitive model, controlling for gene size, variant density, and LD structure, and evaluated against predefined gene sets from Gene Ontology and curated MSigDB pathways (v2023.1.Hs). We tested a total of 17,023 gene sets, and assessed significance after Bonferroni correction (P < 2.94 × 10^-6^).

#### Tissue and cell-type enrichment analyses

For tissue enrichment, we used MAGMA v1.10^89^. Transcriptomic data were obtained from the Human Eye Transcriptome Atlas^66^, which includes gene expression profiles from multiple healthy and diseased eye tissues. Differential expression analysis was used to compute tissue specificity, and the top 10% of genes with the highest tissue-specific t-statistics were selected for each tissue^90^. These were tested for the tissue enrichment using MAGMA’s competitive gene set analysis. Cell type enrichment was performed using ECLIPSER^34^, applied to the 178 genome-wide significant loci identified in the REECAP GWAS.

#### Rare variant association testing

To identify associations between rare coding variants and REECAP embeddings, we computed DeepRVAT^91^ burden scores from exome sequencing data for UKB individuals. We used the same multivariate linear model framework as in the multi-trait GWAS, but testing individual gene-level burden scores rather than individual variants. In total, we assessed 17,984 protein-coding genes.

### Comparison to alternative methods

#### GWAS and colocalization benchmark

We compared multi-trait GWAS (MT-GWAS) discovery using REECAP embeddings to two baseline embeddings: untuned RETFound and ImageNet-pretrained ResNet-50 features, following the TransferGWAS framework^8^. For each method, the first 40 principal components (PCs) were taken as multivariate phenotypes in MT-GWAS. Scalar baselines included cup-to-disc ratio (UKB fields 27857 and 27858) and predicted age, the latter obtained from REECAP embeddings via elastic net leave-one-out cross-validation. Both scalar traits were analyzed using standard single-trait GWAS. All methods were applied to the same subset of UKB fundus images, with left and right eye representations averaged when available. For each method, we identified independent genome-wide significant loci using PLINK clumping (r^2^ < 0.1, 5 Mb window, P < 5 × 10^-8^) and assessed colocalization with ocular disease GWAS from FinnGen using COLOC, as described above.

#### Heritability assessment

SNP-based heritability was estimated for all phenotypes using LD score regression (LDSC)^92^. GWAS summary statistics were first harmonized to HapMap3 variants using `munge_sumstats.py`, then analyzed with `ldsc.py --h2`, using chi-square statistics as the outcome. The analysis employed baselineLD v2.3 LD scores and the corresponding HapMap3 regression weights (`weights_hm3_no_hla`), thereby excluding the extended MHC region and adjusting for broad functional architecture, as recommended by the baseline framework^93,94^. SNP heritability was reported on the observed scale, with standard errors estimated via block jackknife.

#### Polygenic risk score evaluation

We evaluated PRS performance for ocular diseases using genome-wide significant loci (P < 5 × 10⁻⁸) identified via multi-trait GWAS based on REECAP, untuned RETFound, or ImageNet-pretrained ResNet-50 embeddings. Independent variants were selected using LD-based clumping (r^2^ < 0.1, 5 Mb window). Across 20 random 80/20 train/test splits in unrelated UK Biobank participants of European ancestry, we fitted logistic regression models in the training set to predict ICD-10–based ocular disease status (any-time diagnosis) and evaluated AUC in the test set. Method differences were assessed using paired t-tests.

## Data Availability

All data produced in the present study are available upon reasonable request to the authors

## Declarations

### Use of Artificial Intelligence

In the preparation of this manuscript, we utilized the large language model GPT-4 (https://chat.openai.com/) for editing assistance, including language polishing and clarification of text. While this tool assisted in refining the manuscript’s language, it was not used to generate contributions to the original research, data analysis, or interpretation of results. All final content decisions and responsibilities rest with the authors.

## Acknowledgements

We thank Dr. Khalid Aliyev for assistance with the annotation of the reconstructed images, and Brian Clarke for sharing variant annotations used for burden score computations. This research was conducted using the UK Biobank Resource (Application 87065). L.S., E.S., and B.E. were funded by Friedrich-Alexander-University Erlangen-Nuremberg. F.P.C., D.S., and S.C. were funded by the Free State of Bavaria’s High-Tech Agenda through the Institute of AI for Health (AIH). L.S., D.S., and S.C. acknowledge support from HIDSS-006 – Munich School for Data Science@Helmholtz, TUM & LMU. A.V.S. and Q.X. were funded by NIH/NHLBI U01HL166060, and A.V.S. was also supported by NIH/NEI R01 EY031424. J.A.S. acknowledges funding from the Munich Center for Machine Learning (MCML) and the DAAD program Konrad Zuse School of Excellence in Reliable Artificial Intelligence, sponsored by the German Federal Ministry of Research, Technology and Space (BMFTR).

## Authors’ contributions

L.S. implemented the methods and performed the main analyses. D.S., A.A., S.C., and Q.X. contributed to method development and data analysis. A.V.S., J.A.S., E.Sal., and N.C. provided critical guidance on study design, deep learning methodology, and result interpretation. J.S. and B.A. contributed ophthalmological expertise and provided clinical interpretation. O.V.A. and E.Sch. contributed to the integration of mouse clinic resources into the study. L.S. and F.P.C. wrote the manuscript with input from all authors. F.P.C. conceived the project with support from B.E.. F.P.C. and B.E. supervised the study.

