## Supplementary Information for "REECAP: Contrastive learning of retinal aging reveals genetic loci linking morphology to eye disease"

**A2 Supplementary Datasets** 7

**A3 Supplementary Figures** 8

### A1 Supplementary Methods

#### A1.1 REECAP training and representation learning

**REECAP architecture.** We adopted the RETFound foundation model, which is a vision transformer (ViT) backbone pretrained by a masked autoencoder objective on large-scale fundus and OCT images [1]. In RETFound, each input image is divided into non-overlapping patches of size $16 \times 16$, which are linearly projected into token embeddings augmented with positional encodings. The transformer encoder comprises 24 transformer blocks (for the ViT-Large variant) with 16 heads and an embedding dimension of 1024 [2] (i.e. the output embedding size per token is 1024). In REECAP, after passing through the encoder, we apply average pooling over all patch tokens to produce a single fixed-length 1024-dimensional representation vector per image. This image representation is input to the rank and contrast loss.

**RnC loss.** We fine-tuned the RETFound encoder using the Rank-$N$-Contrastive (RnC) objective [3], which enforces that embeddings preserve the ordinal structure of continuous targets—in our case, chronological age. Let a mini-batch contain $B$ individuals with images $\{x_i\}_{i=1}^{B}$ and associated ages $\{y_i\}_{i=1}^{B}$. Each image is augmented twice (see augmentation details below), resulting in $2B$ representations $\{z_\ell\}_{\ell=1}^{2B}$, where $z_\ell \in \mathbb{R}^{1024}$. Define the absolute age distance between samples $i$ and $j$ as

$$d_{i,j} = |y_i - y_j|,$$

and let

$$S_{i,j} = \{\, k \neq i \mid d_{i,k} \geq d_{i,j} \,\}$$

be the set of embeddings that are at least as far in age from anchor $i$ as $j$ is. The per-sample RnC loss is:

$$\ell_i = \frac{1}{2B-1} \sum_{j \neq i} -\log \left( \frac{\exp(-\mathrm{dist}(z_i, z_j)/\tau)}{\sum_{k \in S_{i,j}} \exp(-\mathrm{dist}(z_i, z_k)/\tau)} \right),$$

where $\mathrm{dist}(\cdot, \cdot)$ is Euclidean distance and $\tau$ is a temperature hyperparameter. The total loss is averaged over the batch:

$$\mathcal{L}_{\mathrm{RnC}} = \frac{1}{2B} \sum_{i=1}^{2B} \ell_i.$$

Intuitively, this loss asks: for each anchor embedding, and for each other embedding in the batch, can the model correctly rank that embedding as the most similar among all embeddings whose ages are at least as distant? By minimizing this objective, the model learns an embedding space where proximity reflects age similarity and global ordinal structure is preserved.

**Training details.** We optimized using AdamW (lr $= 5 \times 10^{-6}$, weight decay $= 1 \times 10^{-5}$) with a batch size of 64. We used the following augmentation pipeline:

- Random rotation ($\pm 10°$)
- Random resized crop (90%–100% scale)
- Color jitter (brightness/contrast/saturation $\pm 0.1$, hue $\pm 0.05$)
- Random horizontal flip
- Random perspective distortion (distortion scale 0.05, 50% probability)
- Random affine transforms (translation up to 1% of dims; scale 93%–103%; shear $\pm 2°$)
- Random token (patch) dropout: one patch randomly removed

**Hyperparameter selection and final training.** Hyperparameters were tuned by 5-fold cross-validation across individuals. In each fold:

- 20% of participants formed the held-out "outfold" test set
- The remaining 80% were split into 72% training and 8% validation sets

We tracked metrics (e.g. AUC, correlation) on validation splits to select best hyperparameters. After hyperparameters selection, the model was retrained on the full dataset using the chosen settings. For participants with two fundus images (one per eye), we averaged their two resulting embeddings into a single 1024-dimensional vector. On the cohort level, we then applied PCA and retained the first 40 principal components for downstream analyses.

#### A1.2 Conditional image synthesis

We adopted and extended a framework originally introduced for histology-based GWAS [4] to visualize how scalar traits—such as age, disease status within the next 5 years, or genotype—are reflected in retinal morphology. This method operates on REECAP-derived embeddings from fundus images, enabling both statistical modeling and semantic interpretation of trait-associated phenotypic variation.

1. **Linear Mixed Model.** For a given scalar target variable $\mathbf{t} \in \mathbb{R}^N$ (e.g., genotype dosage, predicted age, or disease label in the next 5 years), we fit a linear mixed model:
$$\mathbf{t} = \mathbf{F}\boldsymbol{\alpha} + \mathbf{X}\boldsymbol{\beta} + \boldsymbol{\varepsilon},$$
where $\mathbf{F} \in \mathbb{R}^{N\times K}$ encodes fixed covariates (e.g., age, sex, genotyping array, genetic PCs), and $\mathbf{X} \in \mathbb{R}^{N\times L}$ contains the retinal embeddings. The embedding-associated effect $\boldsymbol{\beta} \in \mathbb{R}^L$ captures trait-associated morphological variation.

2. **Morphological Effect Axis.** The posterior mean of the random effect vector is computed as:
$$\widehat{\boldsymbol{\beta}} = \hat{\sigma}_X^2 \mathbf{X}^\top \left(\hat{\sigma}_X^2 \mathbf{X}\mathbf{X}^\top + \hat{\sigma}_n^2 \mathbf{I}_N\right)^{-1} (\mathbf{t} - \mathbf{F}\widehat{\boldsymbol{\alpha}}).$$
This defines a morphological effect axis in the embedding space: the direction along which retinal morphology varies with the scalar trait of interest.

3. **Individual-Level Scoring and LOO Estimation.** Each subject's embedding is projected onto this axis to compute an individual-level morphological score:
$$\mathbf{t}_{\mathrm{BLUP}} = \mathbf{X}\widehat{\boldsymbol{\beta}}.$$
To mitigate overfitting, we use the leave-one-out estimate:
$$t_{\mathrm{LOO},i} = \frac{t_{\mathrm{BLUP},i} - H_{ii} \cdot (t_i - \mathbf{F}_i\widehat{\boldsymbol{\alpha}})}{1 - H_{ii}},$$

where $H_{ii}$ is the $i$-th diagonal element of the projection matrix $\mathbf{H} = \hat{\sigma}_X^2 \mathbf{X}\mathbf{X}^\top (\hat{\sigma}_X^2 \mathbf{X}\mathbf{X}^\top + \hat{\sigma}_n^2 \mathbf{I}_N)^{-1}$. These scores are used to stratify individuals by trait-associated morphology.

4. **Latent Embedding Interpolation.** To visualize morphology along the trait axis, we identify representative individuals in the lower and upper tails (e.g., lowest and highest 0.1%) of the LOO score distribution. We compute mean embeddings for each group:

$$\mathbf{z}_{\text{low}} = \text{mean}(\mathbf{X}_{\text{low}}), \quad \mathbf{z}_{\text{high}} = \text{mean}(\mathbf{X}_{\text{high}}),$$

   and linearly interpolate:

$$\mathbf{z}(\alpha) = (1 - \alpha)\, \mathbf{z}_{\text{low}} + \alpha\, \mathbf{z}_{\text{high}}, \quad \alpha \in [0, 1].$$

5. **Image Reconstruction.** While for formulating the LOO predictions we used one 40-dimentional embedding for patient (the result of averaging of embeddings from left and right eye, if both were available), for training the GAN, we have used the 40 first PCs extracted from the representation of the image of each eye separately. Each interpolated embedding $\mathbf{z}(\alpha)$ is decoded using a Progressive GAN trained to reconstruct realistic fundus image conditioning on the embedding, the laterality (if the eye is left or right) and a randomly sampled noise vector. To visualize inter-individual variability, we sample multiple images per interpolation point using different random noise seeds.

   As a specific example, to visualize aging trajectories, we selected the mean embeddings of the youngest and oldest 0.1% of participants (approximately ages 40 and 70), interpolated between them, and decoded three images per step to represent morphological variation across three hypothetical individuals.

All computations scale linearly with the number of individuals, and model fitting is accelerated using low-rank linear algebra identities (e.g., Woodbury identity) and fast reparameterization of variance components [5, 6, 7].

#### A1.3 Multi-trait genome-wide association framework

To identify genetic variants associated with image-derived retinal features, we performed multi-trait GWAS using the REECAP embeddings ( the first $P = 40$ principal components (PCs) of the REECAP representations). Let $\boldsymbol{Y} \in \mathbb{R}^{N \times P}$ denote the matrix of REECAP embeddings for $N$ individuals, $\boldsymbol{g} \in \mathbb{R}^N$ the genotype vector for the tested variant, and $\boldsymbol{F} \in \mathbb{R}^{N \times K}$ the matrix of covariates. The multivariate association model is:

$$\boldsymbol{Y} = \boldsymbol{F}\boldsymbol{A} + \boldsymbol{g}\boldsymbol{b}^\top + \boldsymbol{\Psi},$$

where $\boldsymbol{A} \in \mathbb{R}^{K \times P}$ captures fixed covariate effects, $\boldsymbol{b} \in \mathbb{R}^P$ represents the effect of the variant across embedding dimensions, and $\boldsymbol{\Psi} \in \mathbb{R}^{N \times P}$ is a residual noise matrix. Residuals are assumed to follow a matrix normal distribution with zero mean, independent rows (individuals), and a shared column-wise covariance $\boldsymbol{C} \in \mathbb{R}^{P \times P}$, i.e.,

$\boldsymbol{\Psi} \sim \mathcal{MN}(0, \boldsymbol{I}_N, \boldsymbol{C})$. To improve computational efficiency, we adopted an approximate inference scheme in which the trait covariance $\boldsymbol{C}$ is fully estimated under the null model (excluding genotype effects) and only partially optimized for each variant under the alternative. Briefly, after fitting the null, we rotate the embedding matrix using the eigendecomposition of $\boldsymbol{C}$, transforming the problem into one with uncorrelated phenotypes. In this rotated space, we test for multivariate genetic association by fitting a univariate GWAS model across the decorrelated traits—that is, we optimize a diagonal noise model in the rotated space rather than re-estimating the full covariance. Significance is assessed using a log-likelihood ratio test with $P = 40$ degrees of freedom, comparing the null and alternative models for each variant. $p$-values are computed from the resulting test statistic using a $\chi^2_P$ distribution. Estimated genetic effect sizes in the original embedding space are recovered by inverting the rotation. This approach yields conservative but well-calibrated test statistics and enables efficient genome-wide association testing across millions of variants.

#### A1.4 Summary statistics sources

We utilized genome-wide association summary statistics from the FinnGen consortium, release R12, focusing on ophthalmological and vision-related traits (FinnGen disease group H7: Eye disorders). The following traits were included in our analysis:

- **DM_RETINOPATHY** – Diabetic retinopathy
- **H7_BLINDANDVISIMPAIRMENT** – Blindness and visual impairment
- **H7_MACULADEGEN** – Macular degeneration
- **H7_MODVISIMPBINOC** – Moderate binocular visual impairment
- **H7_RETINALDISOTH** – Other retinal disorders
- **H7_AMD** – Age-related macular degeneration
- **H7_CATARACTSENILE** – Senile cataract
- **H7_MYOPIA** – Myopia
- **H7_GLAUCOMA** – Glaucoma
- **H7_RETINALDETACH** – Retinal detachment

All summary statistics correspond to FinnGen Release 12 (R12) and were obtained from the FinnGen public data repository (`https://www.finngen.fi/en/access_results`).

In addition, we incorporated the following publicly available GWAS summary statistics:

- **Hair colour (natural, pre-greying)** – UK Biobank GWAS summary statistics from the UKB fastGWA imputed phenotype panel (Yang Lab, Westlake University; `https://yanglab.westlake.edu.cn/data/ukb_fastgwa/imp/`).

- **Arterial vascular density** – GWAS summary statistics for image-derived retinal vascular IDPs from Ortín Vela et al. (2024). Data are publicly available via Zenodo (DOI: 10.5281/zenodo.12779552) and the GWAS Catalog.

#### A2 Supplementary Datasets

**Supplementary Dataset 1 — Genome-wide significant loci from REECAP and colocalization results.** Genome-wide significant loci from the multi-trait GWAS of REECAP embeddings ($P < 5 \times 10^{-8}$), including variant-level information, nearest genes, and functional annotations. Additional tabs report colocalization results (using `coloc`) with complex traits and tissues, including FinnGen disease phenotypes, retinal arterial density, and hair color. For each locus–trait pair, posterior probabilities (PP.H0 to PP.H4) are provided, along with indicators of multi-trait colocalization.

**Supplementary Dataset 2 — Top enriched pathways from MAGMA gene-set analysis.** Top 10 enriched pathways identified using MAGMA gene-set analysis via FUMA, based on GWAS summary statistics from REECAP, RETFound, and ImageNet–ResNet-50 embeddings. Each tab corresponds to one model and reports pathway-level results, including number of genes (*N genes*), effect size (Beta), standardized effect size (Beta STD), standard error (SE), raw $P$-value (P), and Bonferroni-adjusted $P$-value (Pbon).

### A3 Supplementary Figures

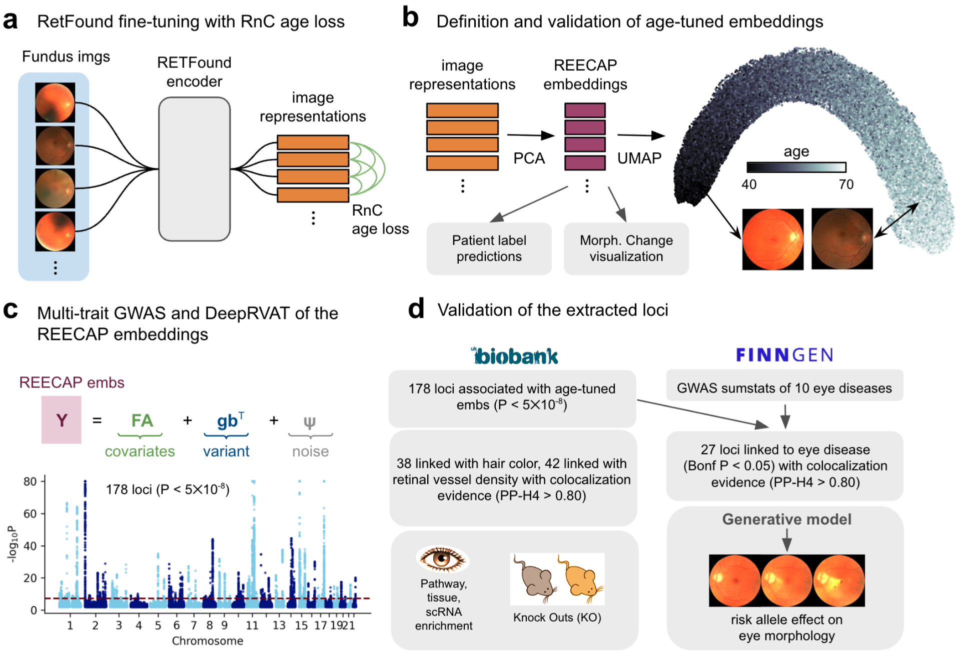

**Supplementary Figure 1 | Overview of the REECAP framework.** (**a**) RETFound encoder fine-tuned using the Rank-$N$-Contrastive (RnC) age loss to derive age-tuned fundus image representations. (**b**) The resulting representations were projected into lower-dimensional REECAP embeddings via PCA and visualized with UMAP, revealing a smooth continuum aligned with chronological age. REECAP embeddings were validated through patient-level prediction tasks and visualization of age- and disease-associated morphological variation. (**c**) Multi-trait GWAS and DeepRVAT analyses on REECAP embeddings identified 178 independent genome-wide significant loci ($P < 5 \times 10^{-8}$) and rare coding variants in seven genes. (**d**) Colocalization and functional validation linked these loci to pigmentation and vascular traits, eye-related pathways, and mouse knockout phenotypes, as well as to ten major ocular diseases from FinnGen. Conditional image synthesis further visualized the effects of selected risk alleles on retinal morphology.

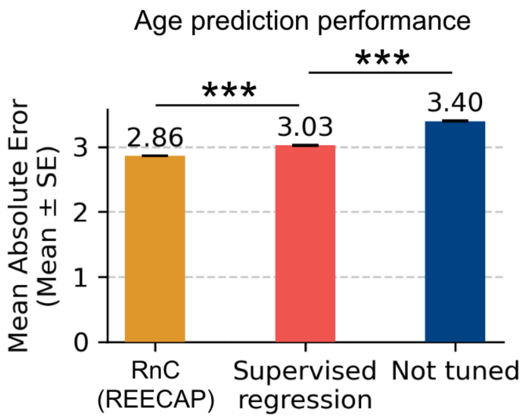

**Supplementary Figure 2 | Performance of contrastive versus supervised age fine-tuning.** Mean absolute error (mean ± s.e., years) for chronological age prediction from fundus image representations. Representations were obtained from the RETFound encoder either fine-tuned using the Rank-$N$-Contrastive (RnC) age loss (REECAP), fine-tuned with conventional supervised regression, or left unmodified (non–age-tuned). Performance was evaluated using 5-fold cross-validation, where embeddings from the held-out fold were used to train and test a linear predictor. RnC fine-tuning achieved the lowest prediction error, significantly outperforming both alternatives (***$P < 10^{-5}$, paired t-test, N=5).

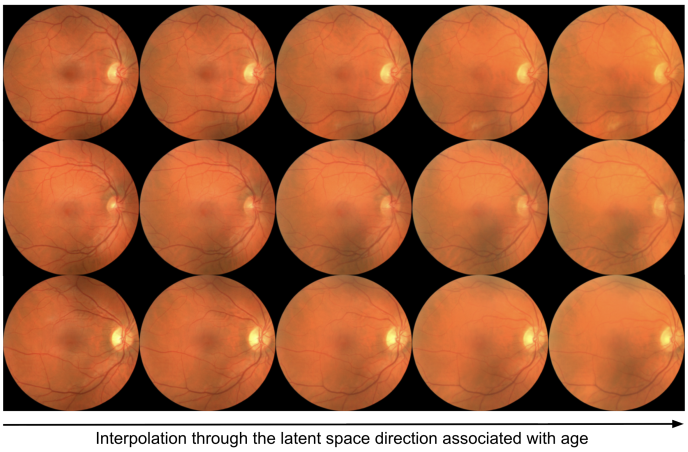

**Supplementary Figure 3 | Synthetic changes along the aging axis in retinal morphology.** Fundus images generated using a Progressive GAN (PGAN) conditioned on the 40 principal components of age-tuned REECAP representations. The leftmost column corresponds to the mean embedding of individuals in the youngest 0.1% of predicted ages (∼40 years), and the rightmost column to the oldest 0.1% (∼70 years). Intermediate columns represent linear interpolations along the age axis in embedding space. Each row shows a different random noise seed to illustrate inter-individual variability. The images reveal synthetic morphological changes consistent with healthy aging, such as progressive vessel narrowing and reduced contrast.

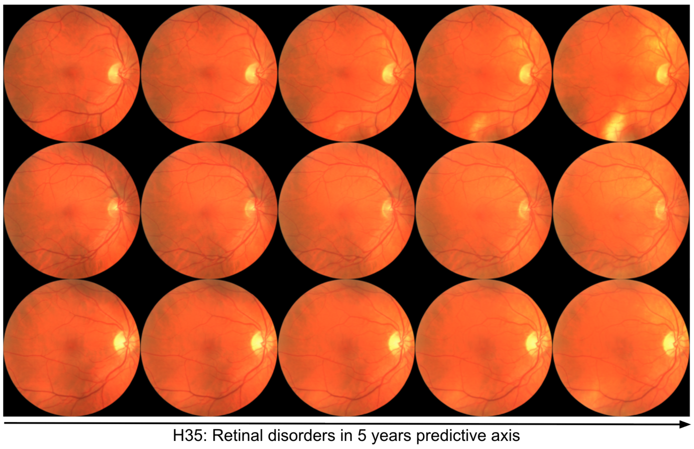

**Supplementary Figure 4 | Synthetic changes along the retinal disorders risk axis.** Fundus images generated using a Progressive GAN (PGAN) conditioned on the first 40 principal components of age-tuned REECAP embeddings. The leftmost column corresponds to the mean embedding of individuals with the lowest predicted risk of retinal disease onset within five years (bottom 0.1% of scores), while the rightmost column corresponds to the highest risk group (top 0.1%). Intermediate columns represent linear interpolations along the risk axis. Each row shows images generated with different noise seeds to reflect inter-individual variability. Features emerging along the risk gradient include central hypopigmentation and pale optic discs.

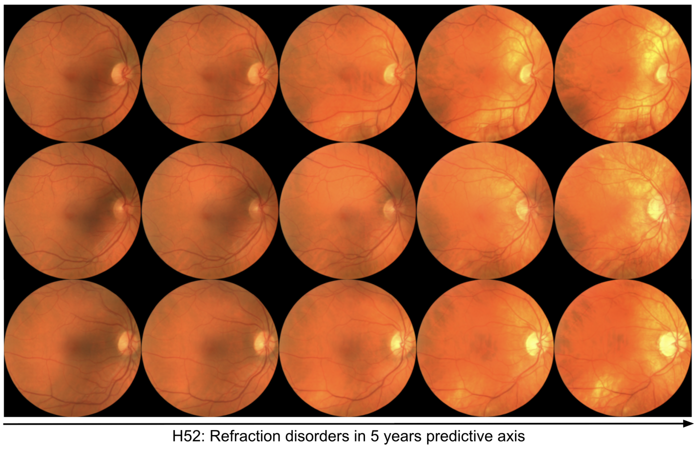

**Supplementary Figure 5 | Synthetic changes along the refraction disorder risk axis.** Analogous to Supplementary Fig. 4, but for predicted risk of refraction disorder within five years. Interpolated images show increasing fundus brightness and an atrophic-appearing, thinner retina with higher risk scores.

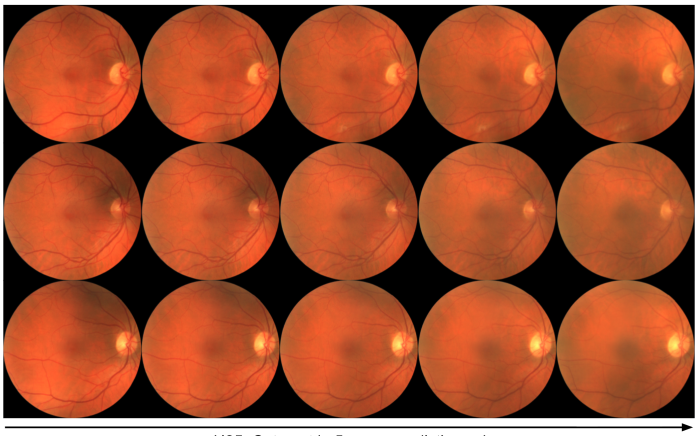

**Supplementary Figure 6 | Synthetic changes along the cataract risk axis.** Analogous to Supplementary Fig. 4, but for predicted risk of cataract within five years. Interpolated images show increased greying, reduced fundus visibility, and subtle vessel thinning associated with higher risk scores.

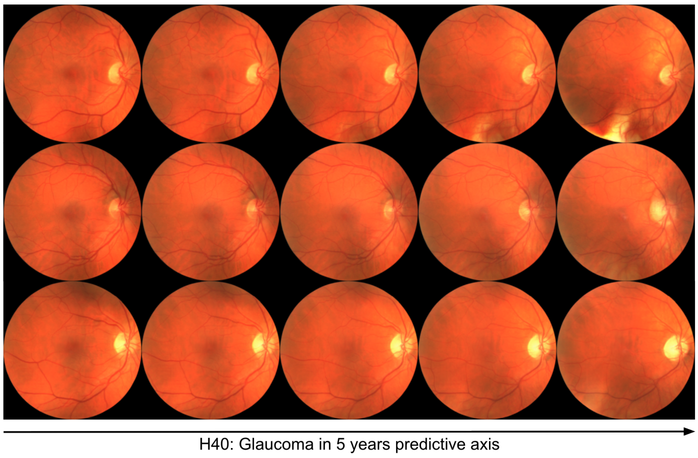

**Supplementary Figure 7 | Synthetic changes along the glaucoma risk axis.** Analogous to Supplementary Fig. 4, but for predicted risk of glaucoma within five years. Interpolated images show increased brighter and a questionably enlarged optic disc with higher risk scores.

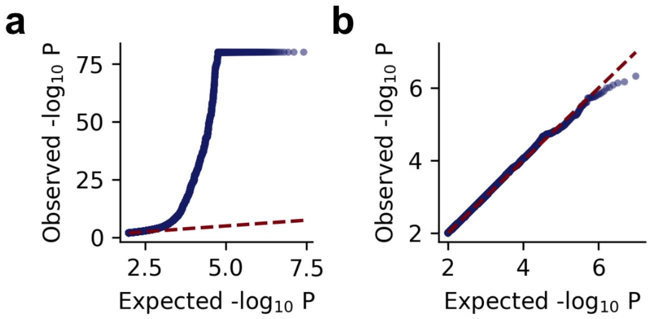

**Supplementary Figure 8 | Quantile–quantile plots for multi-trait GWAS of REECAP embeddings.** QQ plots of REECAP multi-trait GWAS $P$ values for observed data (**a**) and permuted genotypes (**b**).

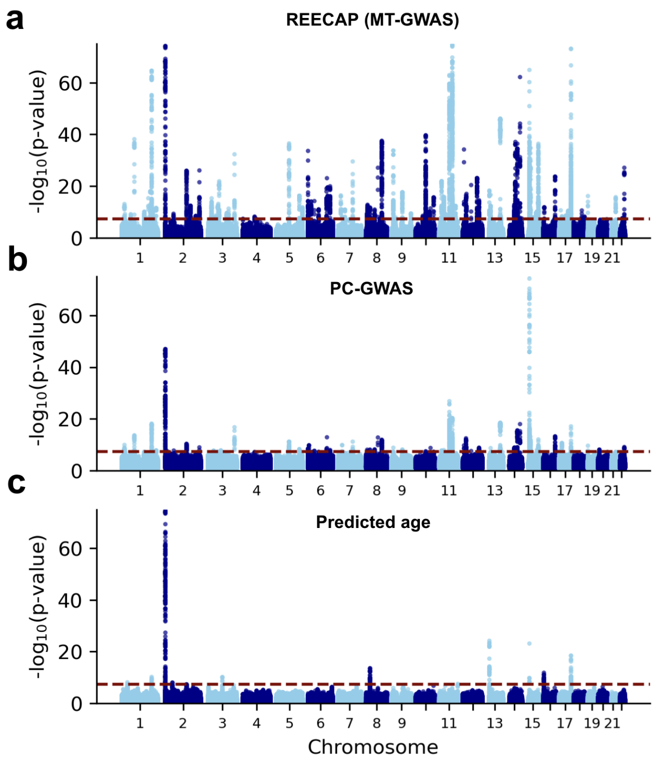

**Supplementary Figure 9 | Manhattan plots comparing multivariate and univariate GWAS strategies on retinal aging traits.** Shown are Manhattan plots of $P$ values from multi-trait GWAS (MT-GWAS) on REECAP embeddings (**a**), univariate GWAS on each principal component of the embeddings with Cauchy-based $P$-value aggregation (**b**), and GWAS on leave-one-out predicted age derived from the embeddings (**c**).

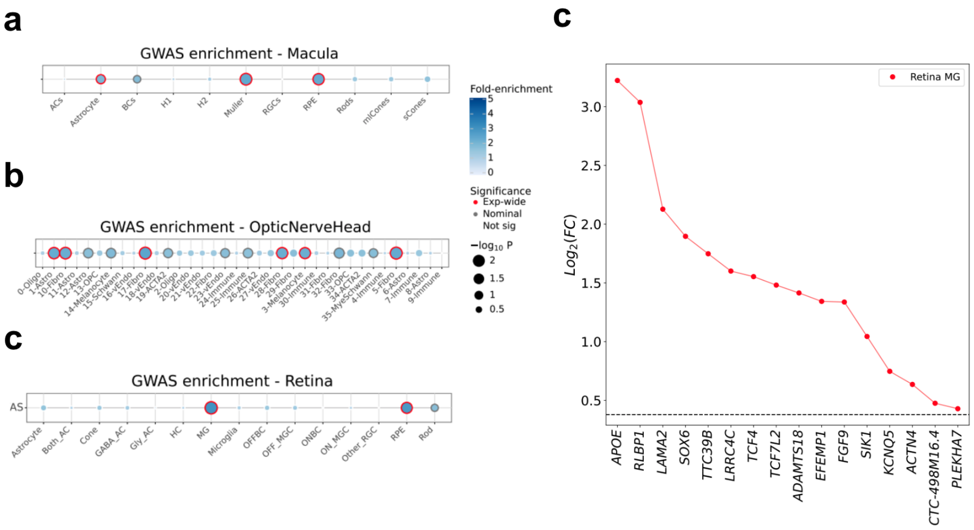

**Supplementary Figure 10 | Cell-type enrichment of REECAP GWAS loci using ECLIPSER.** (**a**–**c**) Cell-type enrichment of 178 independent REECAP GWAS loci in the macula (**a**), optic nerve head (**b**), and retina (**c**). Bubble size represents $-\log_{10}(P)$, color indicates fold-enrichment, and significance is annotated as experiment-wide, nominal, or not significant. (**d**) Top differentially expressed genes enriched in Müller glia from human retina, ranked by $\log_2$ fold change.

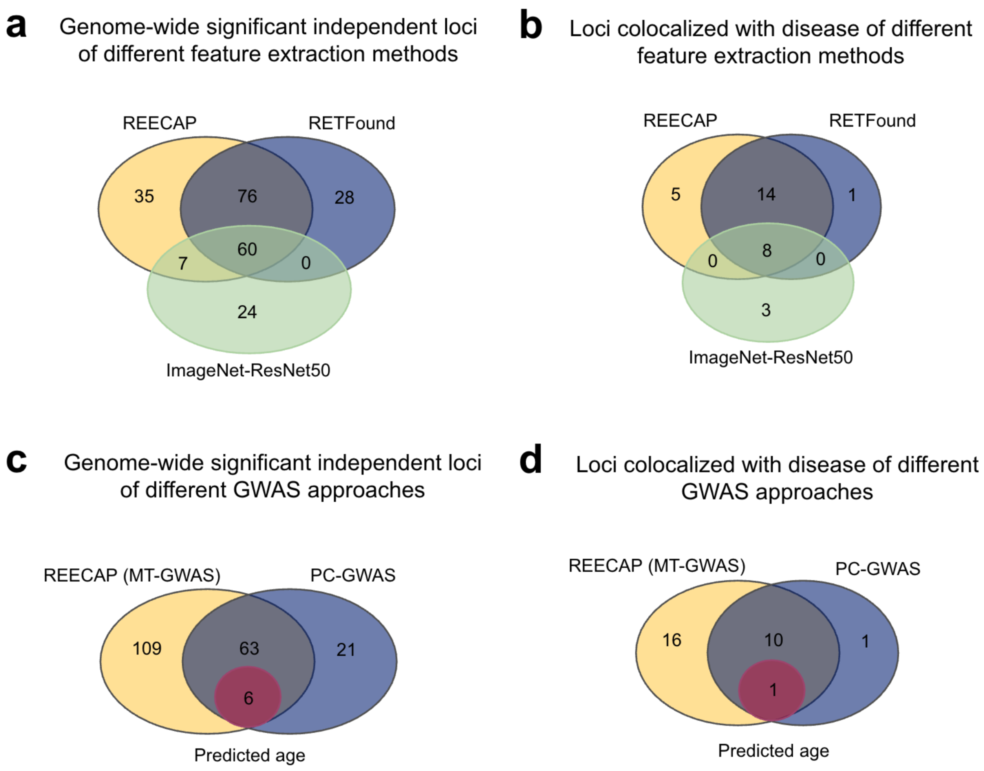

**Supplementary Figure 11 | Comparison of GWAS discovery and disease colocalization across embedding-based and scalar retinal traits.** (**a**) Overlap of independent genome-wide significant loci identified by multi-trait GWAS (MT-GWAS) of REECAP, RETFound, and ImageNet-ResNet50 embeddings. (**b**) Overlap of loci with colocalization evidence ($PP.H4 > 0.8$) with one of 10 FinnGen eye disease GWAS across the same embedding-based approaches. (**c**) Comparison of genome-wide significant loci identified using REECAP MT-GWAS, univariate GWAS on embedding PCs (PC-GWAS), and GWAS of leave-one-out predicted age. (**d**) Overlap of loci with disease colocalization evidence ($PP.H4 \geq 0.8$) among the same three approaches. Lead variants were defined after LD-based clumping ($R^2 < 0.05$ within a 500 kb window) and grouped into loci.

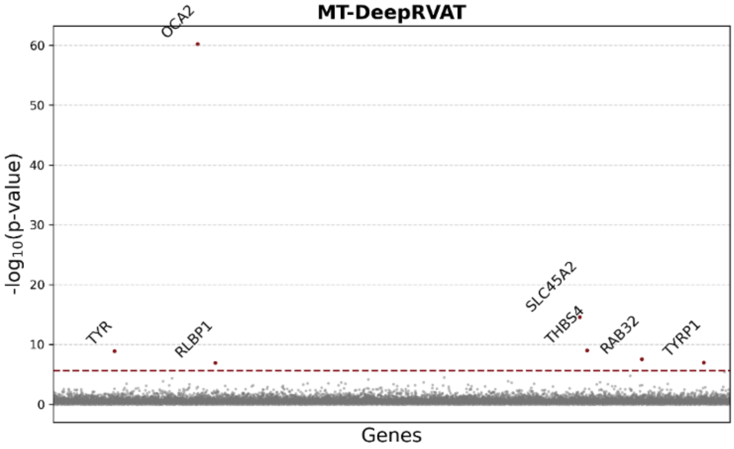

**Supplementary Figure 12 | Manhattan plot of multi-trait rare variant association test of REECAP embeddings.** Shown is the Manhattan plot of $P$ values from multi-trait gene-based burden tests of REECAP embeddings across 17,984 genes in UK Biobank. The red dashed line marks the exome-wide significance threshold.

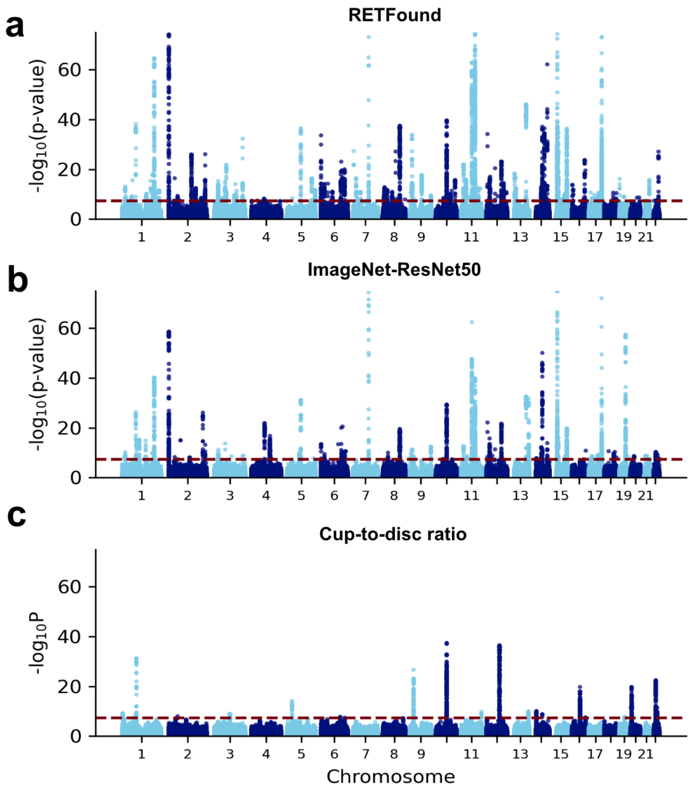

**Supplementary Figure 13 | Manhattan plots comparing multivariate and univariate GWAS strategies on various imaging-derived traits.** Shown are Manhattan plots of $P$ values from multi-trait GWAS (MT-GWAS) on untuned RETFound embeddings (**a**), ImageNet-ResNet50 embeddings (**b**), and univariate GWAS on cup-to-disc ratio (**c**).

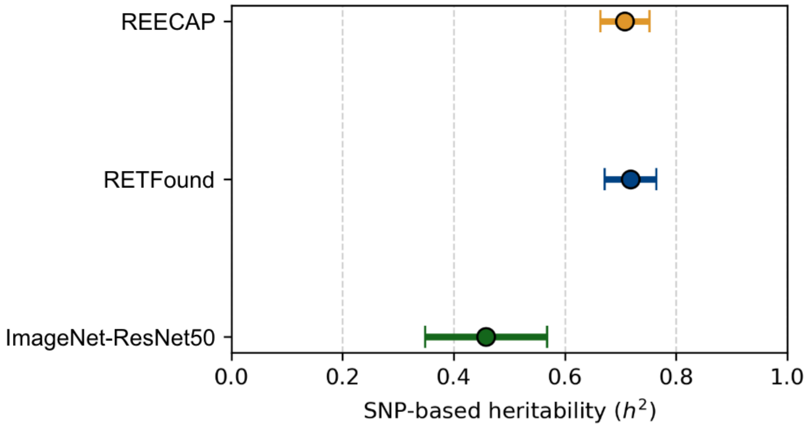

**Supplementary Figure 14 | SNP-based heritability of embedding-derived traits.** SNP-based heritability ($h^2$) estimates from LD Score Regression (LDSC) for REECAP, RETFound, and ImageNet-ResNet50 embeddings. Error bars denote standard errors.

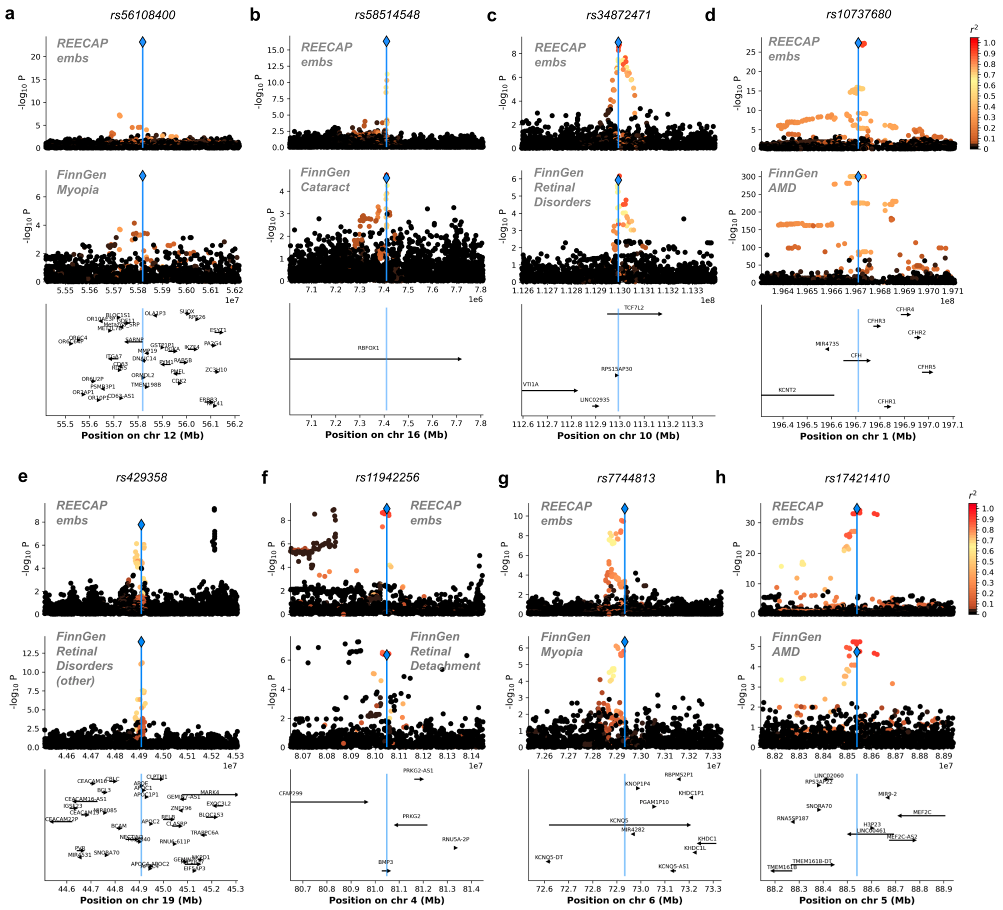

**Supplementary Figure 15 | Genomic and morphological characterization of selected loci from the REECAP GWAS.** (**a**–**h**) Representative loci from the REECAP multi-trait GWAS with evidence of colocalization with eye disease traits (PP-H4 $\geq$ 0.8, see also Supplementary Dataset 1): *rs56108400* near *ORMDL2* (**a**), *rs58514548* in *RBFOX1* (**b**), *rs34872471* in *TCF7L2* (**c**), *rs10737680* in *CFH* (**d**), *rs429358* in *APOE* (**e**), *rs11942256* in *BMP3* (**f**), *rs7744813* in *KCNQ5* (**g**), and *rs17421410* near *LINC00461* (**h**). Each panel shows (from top to bottom): the locus-specific Manhattan plot from the REECAP GWAS, the corresponding FinnGen GWAS for the colocalizing disease trait, and gene annotations in the region.

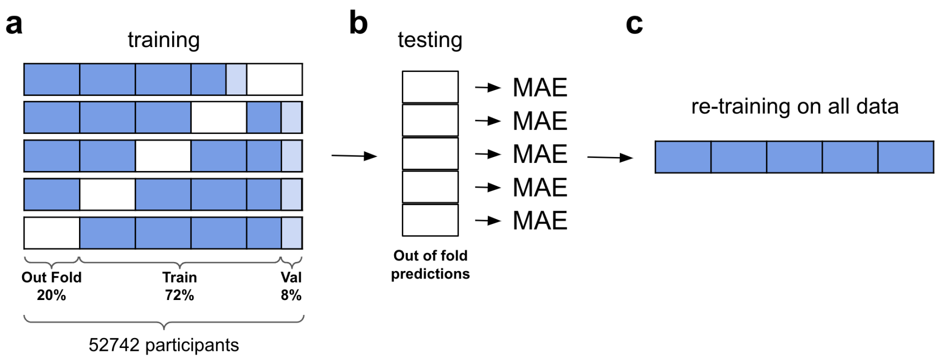

**Supplementary Figure 16 | Cross-validation and model evaluation strategy.** (**a**) Five-fold cross-validation setup used for model development. Each fold includes a split of 72% training, 8% validation (for hyperparameter tuning), and 20% testing (out-of-fold). Participant-level splitting ensures both eyes from the same individual remain in the same fold. (**b**) A ridge regression model is trained on each fold, and mean absolute error (MAE) is computed from out-of-fold predictions. (**c**) The final model is retrained on the full dataset using selected hyperparameters for deployment or downstream analyses.

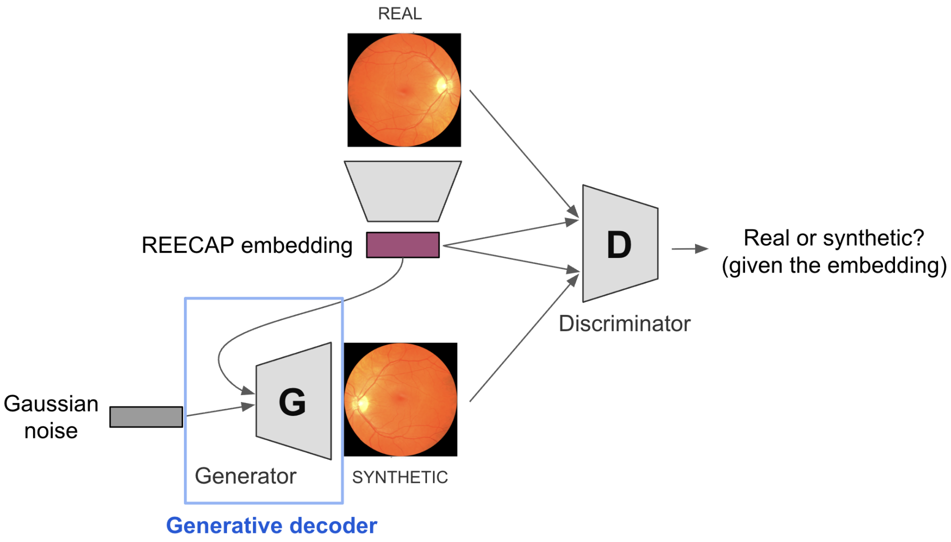

**Supplementary Figure 17 | Conditional GAN framework for synthetic retinal image generation.** Real images are passed through a feature extractor to obtain REECAP embeddings. The generator (G) receives the embedding and Gaussian noise and produces synthetic retinal fundus images. The discriminator (D) learns to distinguish real from synthetic images, conditioned on the embedding. This setup enables image synthesis conditioned on both embedding and noise, disentangling biologically meaningful variation from residual stochasticity. Adapted from HistoGWAS [4].

[3] Kaiwen Zha, Peng Cao, Jeany Son, Yuzhe Yang, and Dina Katabi. Rank-n-contrast: learning continuous representations for regression. Advances in Neural Information Processing Systems, 36, 2024.

[4] Shubham Chaudhary, Almut Voigts, Michael Bereket, Matthew L Albert, Kristina Schwamborn, Eleftheria Zeggini, and Francesco Paolo Casale. Histogwas: An ai-enabled framework for automated genetic analysis of tissue phenotypes in histology cohorts. bioRxiv, pages 2024–06, 2024.

[5] Christoph Lippert, Jennifer Listgarten, Ying Liu, Carl M Kadie, Robert I Davidson, and David Heckerman. Fast linear mixed models for genome-wide association studies. Nature methods, 8(10):833–835, 2011.

[6] Francesco Paolo Casale, Barbara Rakitsch, Christoph Lippert, and Oliver Stegle. Efficient set tests for the genetic analysis of correlated traits. Nature methods, 12 (8):755–758, 2015.

[7] Francesco Paolo Casale. Multivariate linear mixed models for statistical genetics. PhD thesis, 2016.
